# Identifying clinical skill gaps of healthcare workers using a digital clinical decision support algorithm during outpatient pediatric consultations in primary health centers in Rwanda

**DOI:** 10.1101/2025.01.15.25320616

**Authors:** Haykel Karoui, Victor P. Rwandarwacu, Jonathan Niyonzima, Antoinette Makuza, John B. Nkuranga, Valérie D’Acremont, Alexandra V. Kulinkina

**Author notes:** Corresponding author (HK).

## Abstract

Digital clinical decision support algorithms (CDSAs) that guide healthcare workers during consultations can enhance adherence to guidelines and the resulting quality of care. However, this improvement depends on the accuracy of inputs (symptoms and signs) entered by healthcare workers into the digital tool, which relies mainly on their clinical skills, that are often limited, especially in resource-constrained primary care settings. This study aimed to identify and characterize potential clinical skill gaps based on CDSA data patterns and clinical observations. We retrospectively analyzed data from 20,204 pediatric consultations conducted using an IMCI-based CDSA in 16 primary health centers in Rwanda. We focused on clinical signs with numerical values: temperature, mid-upper arm circumference (MUAC), weight, height, z-scores (MUAC for age, weight for age, and weight for height), heart rate, respiratory rate and blood oxygen saturation. Statistical summary measures (frequency of skipped measurements, frequent plausible and implausible values) and their variation in individual health centers compared to the overall average were used to identify 10 health centers with irregular data patterns signaling potential clinical skill gaps. We subsequently observed 188 consultations in these health centers and interviewed healthcare workers to understand potential error causes. Observations indicated basic measurements not being assessed correctly in most children; weight (70%), MUAC (69%), temperature (67%), height (54%). These measures were predominantly conducted by minimally trained non-clinical staff in the registration area. More complex measures, done mostly by healthcare workers in the consultation room, were often skipped: respiratory rate (43%), heart rate (37%), blood oxygen saturation (33%). This was linked to underestimating the importance of these signs in child management, especially in the context of high patient loads typical at primary care level. Addressing clinical skill gaps through in-person training, eLearning and regular personalized mentoring tailored to specific health center needs is imperative to improve quality of care and enhance the benefits of CDSAs.

## Introduction

Children in sub-Saharan Africa experience significant morbidity due to infectious diseases (1,2), with the vast majority of childhood illnesses being addressed at the primary care level. Historically, the quality of primary health services has been poor (3,4). In the 1990s, the United Nations Children’s Fund (UNICEF) and the World Health Organization (WHO) launched the Integrated Management of Childhood Illness (IMCI) initiative. This program aimed to reduce mortality among children under 5 years of age by addressing its five major causes: malaria, diarrhea, measles, malnutrition, and pneumonia (5). While evidence suggests high potential of IMCI to enhance the quality of care and decrease under-five mortality (5–7), its anticipated impact has not been realized. Moreover, for children older than 5 years, who are not covered by IMCI, recommendations are often mixed across various adult guidelines, posing challenges for consistent and effective care.

One of the most significant barriers to the effectiveness of the IMCI program has been poor adherence (8–12). Non-adherence to IMCI guidelines often results in incomplete assessments, with crucial parts omitted (13,14), leading to incorrect classification, misdiagnosis (15), and ultimately, incorrect treatment (16). The reasons for this are manifold, but a key factor is the difficulty healthcare workers (HWs) face in integrating these guidelines into their clinical workflow (16–18). The challenge of adherence is not solely a question of the resources available for adequate and regular training. Even in higher-resource countries like South Africa, the IMCI program has not been able to fulfill its potential due to poor adherence, incomplete assessments, and poor record keeping (19). Therefore, in order for these guidelines to be effective, improving adherence to them among HWs is essential (15).

To address the adherence problem, global health initiatives have shifted their focus from paper-based registers towards digital tools (20,21), such as Clinical Decision Support Algorithms (CDSAs). CDSAs operate by guiding HWs through each step of the outpatient consultation, ultimately proposing a diagnosis and treatment and management plan based on the symptoms, signs, and laboratory test results that have been entered (22). This approach has shown potential in improving adherence to guidelines (9,15,22–25,25–27). However, adherence, conventionally denoting solely the execution of recommended actions, provides an incomplete perspective. In fact, the effectiveness of these tools is heavily dependent on the actual inputs entered by the HWs into the digital platforms, which in turn, are influenced by their clinical skills. In other words, incorrect inputs based on a poor-quality history taking and physical exam may still result in incorrect diagnosis and treatment being proposed. There is a lack of studies focusing on the quality of clinical skills and child assessments done by HWs at the primary care level using CDSAs. This represents a significant gap in our understanding of the effectiveness of these digital tools, and of IMCI in general.

The DYNAMIC project, a five-year research study initiated in 2021 in the Western Province of Rwanda, aimed to implement and validate ePOCT+ (electronic point of care tests plus), a digital IMCI-based CDSA created to improve the quality of outpatient care for children under 15 years of age. As evidenced by previous studies, increased adherence to guidelines was observed; however, clinical outcomes did not improve. This realization prompted a deeper investigation, utilizing the data from the CDSA, to explore clinical skills and assess the quality of HW assessments beyond the conventional evaluation of adherence. We sought to not only determine if assessments were performed, but also to evaluate the accuracy and thoroughness with which they were conducted. This exploratory study aimed to identify and characterize potential clinical skill gaps based on ePOCT+ data patterns and clinical observations. The findings of this study provide valuable insights into the role of HWs’ clinical skills in the effective use of CDSAs like ePOCT+.

## Methodology

### Study context

ePOCT+ was implemented in 16 primary healthcare centers (HCs) of Rusizi and Nyamasheke districts in Rwanda. While ePOCT+ is built on IMCI as a backbone, it considers a broader range of diagnoses to accommodate a wider pediatric age range (1 day to 14 years) and utilizes point-of-care devices and tests, such as pulse oximetry, hemoglobin level, and C-reactive protein measurement (28). The primary aim of ePOCT+ is to guide HWs throughout the consultation by streamlining the diagnostic process and suggesting appropriate management strategies. To achieve this, HWs input various information into a tablet-based application following a logical consultation flow, including anthropometric measures, vital signs, and the presence or absence of specific symptoms and/or signs based on the patient’s complaints ((28,29); the algorithm then proposes certain diagnostic tests. Finally, clinical diagnoses based on these inputs are proposed, which can be either accepted or rejected by the HW, along with the corresponding guidelines for managing those diseases, including detailed treatment information. The intended use of the algorithm is in real-time, i.e., HWs are supposed to do an assessment (e.g., measure the respiratory rate) and input the result, then move on to the next proposed task. All inputs are mandatory; however, in certain situations, such as when time is limited or the patient is unstable or agitated, HWs are given the option of marking a select few assessments as “not feasible”. While it is discouraged, HWs may decide to answer questions without conducting the assessment, such as assuming that the child does not have conjunctival pallor without checking. Because HWs are most likely to enter negative responses in such cases, we expect that the dataset may be biased towards negative responses, especially in the case of binary variables indicating presence or absence of a condition. Additionally, some health centers hire staff with or without clinical background and capacitate them to take certain anthropometric measurements in the reception area before the child’s consultation with the HWs.

This study utilized a mixed methods approach to identify and characterize potential clinical skill gaps in HCs. First, a retrospective analysis of ePOCT+ data was conducted to detect unusual patterns reflecting potential clinical skill gaps. Subsequently, consultations were prospectively observed and HWs interviewed, to further explore and try to understand these gaps.

### Retrospective analysis

#### Data sources

ePOCT+ stores all the information (date of consultation, anthropometric measures, vitals, presence/absence of specific symptoms and signs prompted by the algorithm, diagnoses, medicines, managements, etc.) entered by the HW in the tablet during consultations. We retrospectively analyzed data from 20,204 outpatient consultations conducted between November 2021 and October 2022 with children aged 1 day to 14 years with an acute condition, in the 16 HCs where the intervention was deployed. The data were accessed for research purposes on 22 November 2022. All data used in the study were anonymized, and the authors did not have access to information that could identify individual participants during or after data collection. Data cleaning, management, and analyses were conducted using R software (version 4.2.1).

#### Signs of interest

Most of the signs in ePOCT+ are binary (presence/absence), which can be challenging to ground-truth (and are potentially biased toward negative responses as described above). On the other hand, some of the clinical signs, particularly measurements that result in numerical values, can be observed in a more objective and offer informative distributions for detecting outliers and other meaningful patterns. Thus, we focused on quantifiable and objective measures: temperature, mid-upper arm circumference (MUAC), weight, height, MUAC for age z-score, weight for age z-score, weight for height z-score, heart rate, respiratory rate and blood oxygen saturation (SpO_2_). Z-scores were considered because MUAC, weight, and height distributions are age-specific and require normalization to be meaningful. The way each sign is measured and recorded by the HWs (or automatically calculated in the application such as z-scores) is described in Table 1.

**Table 1:** Summary of signs of interest, how they were assessed by healthcare workers, entered in the tablet, and subsequently analyzed.

| Sign | Material/tool used for assessment | Performed in all or a subset of children | Data format | Options available in the CDSA if measurement is not performed | Statistical analyses of interest |
| --- | --- | --- | --- | --- | --- |
| Temperature | - Infrared thermometer<br>- Axillary digital thermometer<br>- Axillary mercury thermometer | All | Degrees Celsius in decimal format<br>(XX.X °C) | Can tick "Not feasible" (becomes missing); in this case, a subsequent question asks if the child "feels hot" or "does not feel hot". | - % of most frequent values<br>- % missing values<br>- Mean |
| MUAC | - UNICEF MUAC tape | Age > 6 months | Centimeter in decimal format<br>(XX. X cm) | Can tick "Not feasible" (considered as missing). | - % of most frequent values<br>- % of missing values |
| Weight | - Adult balance<br>- Neonatal balance | All | Kilograms in decimal format<br>(XX.X kg) | Can be marked as either "measured" or "estimated". Only "measured" values are considered in the present analysis, "estimated" values are considered as missing. | - % of most frequent values<br>- % missing values |
| Height | - UNICEF Length board<br>- Adult length meter | All | Centimeter in decimal format<br>(XXX.X cm) | Can tick "Not feasible" (considered as missing). | - % of most frequent values<br>- % of missing values |
| MUAC for age z-score | Background Calculation (Based on WHO child growth standard) | Age > 6 months | Integer format with values from -3 to 3 (X) | Not calculated | - % of extreme values (<-2 or >2) |
| Weight for age z-score | Background Calculation (Based on WHO child growth standard) | All | Integer format with values from -3 to 3 (X) | Not calculated | - % of extreme values (<-2 or >2) |
| Weight for height z-score | Background Calculation (Based on WHO child growth standard) | All | Integer format with values from -3 to 3 (X) | Not calculated | - % of extreme values (<-2 or >2) |
| Heart rate | - Pulse Oximeter (with neonatal, child, adult probes) | Only children with a Respiratory main complaint | Heart beats per minute in whole number<br>(XXX beats/minute) | Can tick "Not feasible" (considered as missing). | - % of most frequent value<br>- % of missing value<br>- Mean<br>- IQR |
| SpO<sub>2</sub> | - Pulse Oximeter (with neonatal, child, adult probes) | Only children with a Respiratory main complaint | Whole percentage format<br>(X%) | Can tick "Not feasible" (considered as missing). | - % of most frequent value<br>- % of missing value<br>- Mean<br>- IQR |
| Respiratory rate | - Timer<br>- RRate 3.3 Android app | Only children with a Respiratory main complaint | Breaths per minute in whole number format<br>(XX breaths/minute) | Can tick "Not feasible" (considered as missing); a subsequent question asks if it is "visibly fast" or "visibly normal". | - % of most frequent values<br>- % of missing values<br>- Mean<br>- IQR |

IQR = inter-quartile range; MUAC = mid-upper arm circumference; SpO<sub>2</sub> = Blood oxygen saturation

For all the signs of interest, we identified the most frequent value (the value and its frequency) and the percent of measures that was declared as not feasible by the HW and marked as a missing value in the dataset. For the anthropometric signs (MUAC, weight, height) we also considered the percent of extreme values (<-2 or >2) in the z-scores. For temperature, blood oxygen saturation, heart rate and respiratory rate, we considered the median and the inter-quartile range (IQR).

#### Identification of potential skill gaps

We compared the statistical parameters for each sign of interest in individual HCs against the same parameter across all HCs (Table 2), which we considered as our reference due to a lack of a true gold standard. We conducted the analysis at HC level, allowing us to identify the HCs that exhibited most significant deviations, guiding further investigation in subsequent analyses. Based on the features of the statistical parameters, signs (per HC) were labeled with one of the four distinct pattern types (i.e., skipper, mono-repeater, multi-repeater, and wrongly evaluated), along with one of the three corresponding levels of expression (i.e., mild, moderate, or severe). The level of expression was determined by comparing the statistical parameters (mean, interquartile range [IQR], frequency of not feasible/missing values, and value distribution) of clinical measurements across different HCs with the combined average from all HCs. The degree of deviation from this combined baseline was then categorized as mild, moderate, or severe, depending on how much the values diverged, as illustrated in Table 2. Examples of how data were visualized, and patterns were identified are provided in Figs S1-S5 and Table S2, Supporting Information.

**Table 2:** Types of pattern and definitions for their level of deviation from the mean values and distributions for all health centers (considered as reference).

| Type of values pattern | Definition | Mild | Moderate | Severe |
| --- | --- | --- | --- | --- |
| Skipper | High frequency of “not feasible” and thus missing values.<br>*[does not apply to temperature, weight, and z-scores]* | 5-10% more frequent compared to reference* | 10-30% more frequent compared to reference | >30% more frequent compared to reference |
| Mono-repeater | High percentage of one value or of rounded values; a small IQR** with the distribution of values being concentrated around one unique value.<br>*[does not apply to weight, height, and z-scores]* | The value is 5-10% more frequent than the reference | The values are 10-30% more frequent than the reference | The values are >30% more frequent than the reference |
|  |  | 25-50% smaller IQR* than reference | 50-100% smaller IQR* than reference | >100% smaller IQR* than reference |
| Multi-repeater | High percentage of 2 to 3 values or round values with the distribution of values being concentrated around 2 to 3 values.<br>*[does not apply to weight, height, and z-scores]* | The values are 5-10% more frequent than the reference | The values are 10-30% more frequent than the reference | The values are >30% more frequent than the reference |
|  |  | 25-50% smaller IQR* than reference | 50-100% smaller IQR* than reference | >100% smaller IQR* than reference |
| Wrongly evaluated | High percentage of outlier values, indicating potential errors in measurements (e.g., high level of extreme z-score values, or implausible mean of values (e.g., mean temperature of 41°C))<br>*[does not apply to weight and height]* | Implausible mean of values | 10-30% more outlier values in z-scores | >30% more outlier values in z-scores |
|  |  | 5-10% more outlier values in z-scores |  |  |

\*IQR: inter-quartile range

### Prospective observation of consultations

Based on the results of the retrospective analysis, we observed 188 routine consultations from 09 January 2023 and to 09 March 2023 in a subset of 10 of 16 HCs (approximately 19 observations per HC). The selection of HCs was guided by the retrospective analysis, ensuring that the 10 HCs chosen were those showing the most critical results. The overall sample size allowed for sufficient variety while maximizing available time and resources for the study. The observing study clinician obtained oral consent from the HWs, which was documented and witnessed by a DYNAMIC field team member, and was instructed not to interfere with the consultation to avoid introducing any additional bias to the observer effect. To ensure a standardized and consistent evaluation, a digital evaluation form (Google sheets) was used (Table S3). The same signs and symptoms were evaluated with observations as those studied in the retrospective analysis above. These observations were conducted over 3 days per HC, with efforts made to separate them by a few days in order to have more chance to observe several different HWs and minimize potential bias. This separation aimed to reduce the likelihood that HWs would alter their behavior after the interviews conducted at the end of each consultation day, ensuring more natural and representative observations of their clinical practices. Most of the time, there was only one HW attending to children in the HC on a given day. On the rare occasions when two HW were present, each was observed by one of the two study clinicians. At the end of each day of observation in a HC (and not after each consultation to avoid any influence on subsequent consultations), the observing study clinician conducted an interview with the HW to understand why the assessment of some signs was skipped. For each measure of interest, the study clinician documented whether it was done or skipped, and the quality of the assessment as sufficient or insufficient. Additionally, explanations provided by HWs regarding measures being skipped or incorrectly performed were also recorded. Data were exported to Microsoft Excel (Version 16.77.1) for further simple descriptive analysis.

## Results

### Retrospective data analysis

Variability in the quality of assessment was observed across clinical signs and HCs (Table 3). Some signs exhibited a consistent pattern in all HCs (e.g., height and the three z-scores) whereas others exhibited diverse patterns (e.g., MUAC, HR, RR). Most HCs had multiple issues with the studied measurements, whereas some had very few or none at all. The Supporting Information section provide detailed visualizations and distributions: Table S2 shows the distribution of the “not feasible” status of each sign stratified by HF, Fig S3 presents boxplots of each sign stratified by HF, Fig S4 displays density plots of each sign stratified by HF, and Fig S5 illustrates Z-scores distribution plots for each sign stratified by HF. Based on the summary (Table 3), we selected the 10 lowest performing HCs for observation. The criteria for the selection of these HCs included the presence of at least two signs whose values had a pattern showing deviation, with at least one pattern exhibiting a level of deviation categorized as moderate or severe.

**Table 3:** Summary of data patterns and indicative behavior classifications.

| Health Centers | Temperature | MUAC | Weight | Height | MUAC for age* | Weight for age* | Weight for height* | HR | SpO<sub>2</sub> | RR | Selected for observation |
| --- | --- | --- | --- | --- | --- | --- | --- | --- | --- | --- | --- |
| HC 1-1 | - | - | - | - | - | - | - | - | - | Mono - WE | No |
| HC 1-2 | - | - | - | - | WE | - | - | Skipper | Skipper - Mono | Multi | Yes |
| HC 1-3 | - | Multi | - | - | - | - | - | Skipper - Multi | Skipper - Mono | Skipper - Mono | Yes |
| HC 1-4 | - | - | - | Skipper | - | - | WE | - | - | - | Yes |
| HC 1-5 | - | Multi | - | Skipper | - | - | WE | Multi | - | - | Yes |
| HC 2-1 | - | - | - | - | - | - | - | - | - | - | No |
| HC 2-2 | - | Skipper | - | Skipper | WE | - | WE | Mono | - | - | Yes |
| HC 2-3 | - | Skipper - Multi | - | Skipper | - | - | - | - | - | WE | Yes |
| HC 2-4 | Multi | - | - | - | - | WE | WE | - | - | Mono | Yes |
| HC 2-5 | Mono | - | - | - | - | - | - | Skipper | Multi | Multi | Yes |
| HC 3-1 | - | - | - | - | - | - | - | - | Multi | - | No |
| HC 3-2 | - | - | - | - | - | - | - | - | Multi | WE | No |
| HC 3-3 | - | - | - | - | - | - | - | - | - | - | No |
| HC 3-4 | - | - | - | - | - | - | - | Mono | - | - | No |
| HC 3-5 | - | Skipper - Mono | - | Skipper | WE | WE | WE | Skipper | Skipper - Mono | Skipper | Yes |
| HC 3-6 | Multi | Skipper - Mono | - | Skipper | WE | - | WE | Skipper - Mono | Skipper - WE | Skipper - Mono | Yes |

\*Z-scores; MUAC = mid-upper arm circumference; HR = heart rate; SpO<sub>2</sub> = Blood oxygen saturation; RR = respiratory rate
- Level of pattern deviation: No deviation [ - ] ; Mild ; Moderate ; Severe .
- Definition of sign values patterns (compared to the means and distributions for all health centers): Skipper: More missing/not feasible values; Mono (mono-repeater): High frequency of one value; Multi (multi-repeater): Multiple values with high frequency and limited variation; WE (Wrongly evaluated): unexpected data patterns/types or outlier values.
- Location of measurement of the sign within the health center: Done by somebody else than the HW in charge, before the consultation ; Done by HW in consultation

### Prospective observation of consultations

According to the prospective observations (Fig 1), respiratory rate, heart rate, and blood oxygen saturation were the most frequently skipped (43%, 37%, and 33%, respectively). Temperature, weight, and MUAC were more often assessed, but with insufficient quality (67%, 70%, and 69%, respectively). Height showed a mixed pattern, with 21% of assessments skipped and 54% deemed insufficient.

**Fig 1:** Distribution of the performance and quality of assessment for each sign, as observed by the study clinician. When using the CDSA, some signs are proposed to be assessed only in sub-groups of patients reporting specific main complaints, resulting in different numbers of consultations observed according to the sign. Temperature, weight, and height were asked for all children (188 consultations each). MUAC was asked only for children aged > 6 months. Respiratory rate, heart rate, and blood oxygen saturation depended on specific complaints.

The most common reason for skipping measurements was that the sign was not deemed necessary based on the child’s symptoms. This was particularly frequent for heart rate (67%), respiratory rate (60%), blood oxygen saturation (59%), weight (56%), and MUAC (52%) (Table 4). Time constraints also played a significant role, especially for heart rate (52%) and blood oxygen saturation (41%), where the assessment was perceived as too time-consuming. Child-related factors, such as agitation, also contributed to skipped assessments, particularly for blood oxygen saturation (15%) and respiratory rate (10%). For measurements that were performed but deemed of insufficient quality, the most frequent issues were related to poor methodology. This included improper band placement for MUAC (97%), incorrect use of thermometers for temperature (87%), and improper positioning for height (77%). Equipment-related problems were also prevalent, especially for blood oxygen saturation and heart rate, where inadequate oximeter sensors for the child’s age were used in 56% of cases. Additionally, interference from clothing or footwear significantly impacted the accuracy of weight (89%) and height (61%) measurements.

**Table 4:**
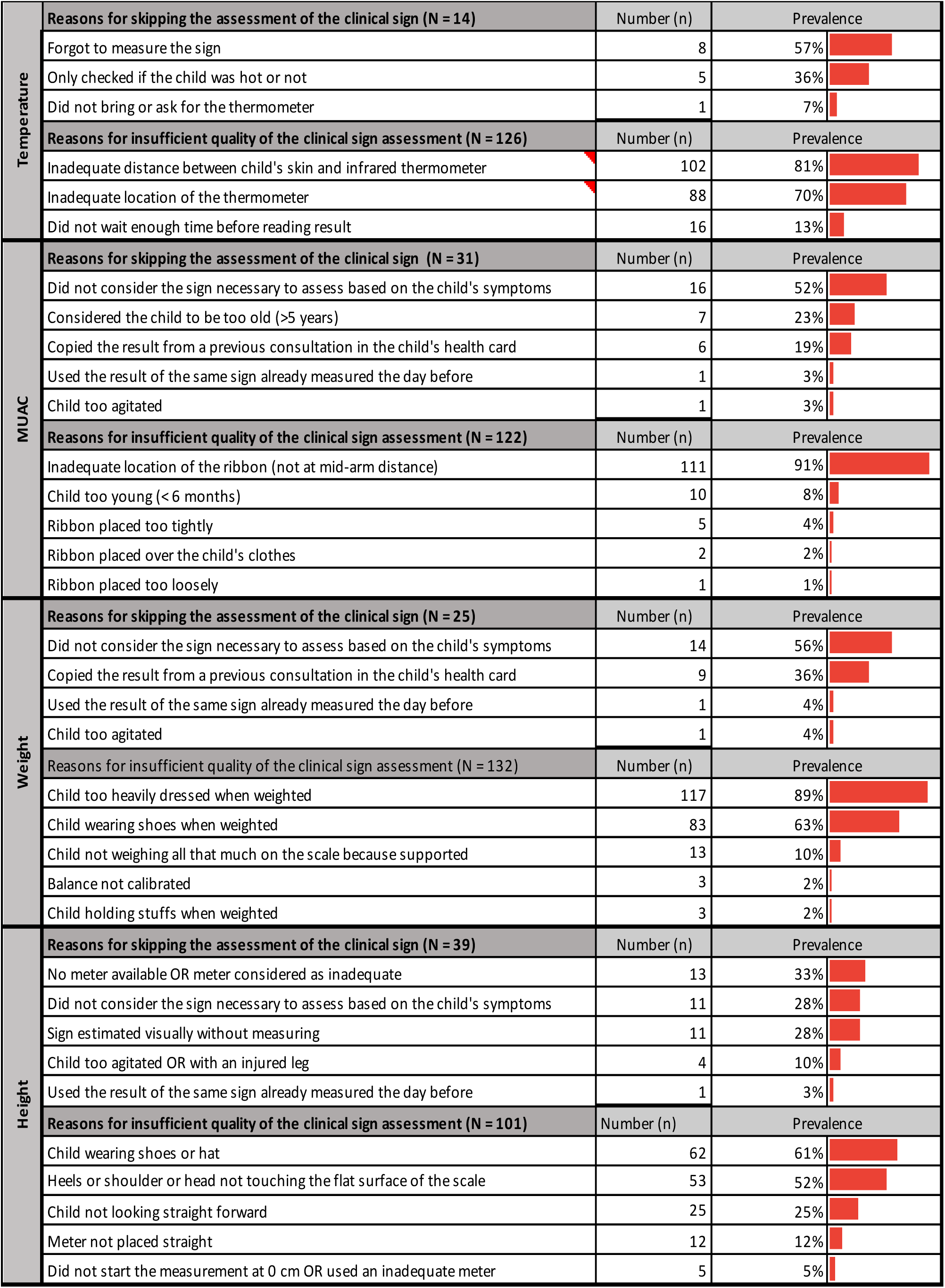
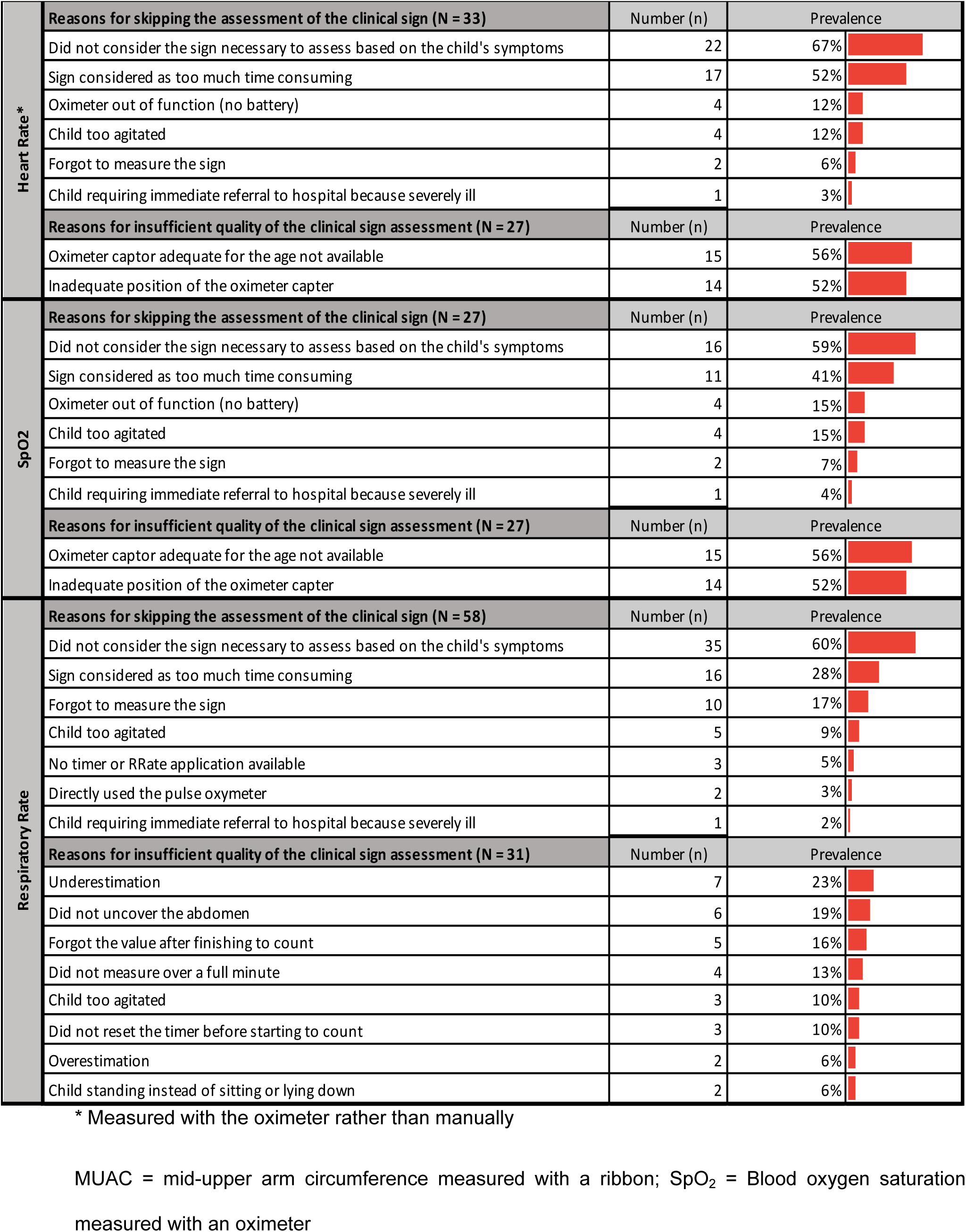
Reasons provided by routine HWs for skipping a sign, or for insufficient quality of its measurement.

|  |  |  |  |
| --- | --- | --- | --- |
| Temperature | **Reasons for skipping the assessment of the clinical sign (N = 14)** | Number (n) | Prevalence |
|  | Forgot to measure the sign | 8 | 57% |
|  | Only checked if the child was hot or not | 5 | 36% |
|  | Did not bring or ask for the thermometer | 1 | 7% |
|  | **Reasons for insufficient quality of the clinical sign assessment (N = 126)** | Number (n) | Prevalence |
|  | Inadequate distance between child's skin and infrared thermometer | 102 | 81% |
|  | Inadequate location of the thermometer | 88 | 70% |
|  | Did not wait enough time before reading result | 16 | 13% |
| MUAC | **Reasons for skipping the assessment of the clinical sign (N = 31)** | Number (n) | Prevalence |
|  | Did not consider the sign necessary to assess based on the child's symptoms | 16 | 52% |
|  | Considered the child to be too old (>5 years) | 7 | 23% |
|  | Copied the result from a previous consultation in the child's health card | 6 | 19% |
|  | Used the result of the same sign already measured the day before | 1 | 3% |
|  | Child too agitated | 1 | 3% |
|  | **Reasons for insufficient quality of the clinical sign assessment (N = 122)** | Number (n) | Prevalence |
|  | Inadequate location of the ribbon (not at mid-arm distance) | 111 | 91% |
|  | Child too young (< 6 months) | 10 | 8% |
|  | Ribbon placed too tightly | 5 | 4% |
|  | Ribbon placed over the child's clothes | 2 | 2% |
|  | Ribbon placed too loosely | 1 | 1% |
| Weight | **Reasons for skipping the assessment of the clinical sign (N = 25)** | Number (n) | Prevalence |
|  | Did not consider the sign necessary to assess based on the child's symptoms | 14 | 56% |
|  | Copied the result from a previous consultation in the child's health card | 9 | 36% |
|  | Used the result of the same sign already measured the day before | 1 | 4% |
|  | Child too agitated | 1 | 4% |
|  | **Reasons for insufficient quality of the clinical sign assessment (N = 132)** | Number (n) | Prevalence |
|  | Child too heavily dressed when weighted | 117 | 89% |
|  | Child wearing shoes when weighted | 83 | 63% |
|  | Child not weighing all that much on the scale because supported | 13 | 10% |
|  | Balance not calibrated | 3 | 2% |
|  | Child holding stuffs when weighted | 3 | 2% |
| Height | **Reasons for skipping the assessment of the clinical sign (N = 39)** | Number (n) | Prevalence |
|  | No meter available OR meter considered as inadequate | 13 | 33% |
|  | Did not consider the sign necessary to assess based on the child's symptoms | 11 | 28% |
|  | Sign estimated visually without measuring | 11 | 28% |
|  | Child too agitated OR with an injured leg | 4 | 10% |
|  | Used the result of the same sign already measured the day before | 1 | 3% |
|  | **Reasons for insufficient quality of the clinical sign assessment (N = 101)** | Number (n) | Prevalence |
|  | Child wearing shoes or hat | 62 | 61% |
|  | Heels or shoulder or head not touching the flat surface of the scale | 53 | 52% |
|  | Child not looking straight forward | 25 | 25% |
|  | Meter not placed straight | 12 | 12% |
|  | Did not start the measurement at 0 cm OR used an inadequate meter | 5 | 5% |

| Heart Rate * | Reasons for skipping the assessment of the clinical sign (N = 33) | Number (n) | Prevalence |
| --- | --- | --- | --- |
|  | Did not consider the sign necessary to assess based on the child's symptoms | 22 | 67% |
|  | Sign considered as too much time consuming | 17 | 52% |
|  | Oximeter out of function (no battery) | 4 | 12% |
|  | Child too agitated | 4 | 12% |
|  | Forgot to measure the sign | 2 | 6% |
|  | Child requiring immediate referral to hospital because severely ill | 1 | 3% |
|  | Reasons for insufficient quality of the clinical sign assessment (N = 27) | Number (n) | Prevalence |
|  | Oximeter captor adequate for the age not available | 15 | 56% |
|  | Inadequate position of the oximeter capter | 14 | 52% |
| SpO2 | Reasons for skipping the assessment of the clinical sign (N = 27) | Number (n) | Prevalence |
|  | Did not consider the sign necessary to assess based on the child's symptoms | 16 | 59% |
|  | Sign considered as too much time consuming | 11 | 41% |
|  | Oximeter out of function (no battery) | 4 | 15% |
|  | Child too agitated | 4 | 15% |
|  | Forgot to measure the sign | 2 | 7% |
|  | Child requiring immediate referral to hospital because severely ill | 1 | 4% |
|  | Reasons for insufficient quality of the clinical sign assessment (N = 27) | Number (n) | Prevalence |
|  | Oximeter captor adequate for the age not available | 15 | 56% |
|  | Inadequate position of the oximeter capter | 14 | 52% |
| Respiratory Rate | Reasons for skipping the assessment of the clinical sign (N = 58) | Number (n) | Prevalence |
|  | Did not consider the sign necessary to assess based on the child's symptoms | 35 | 60% |
|  | Sign considered as too much time consuming | 16 | 28% |
|  | Forgot to measure the sign | 10 | 17% |
|  | Child too agitated | 5 | 9% |
|  | No timer or RRate application available | 3 | 5% |
|  | Directly used the pulse oxymeter | 2 | 3% |
|  | Child requiring immediate referral to hospital because severely ill | 1 | 2% |
|  | Reasons for insufficient quality of the clinical sign assessment (N = 31) | Number (n) | Prevalence |
|  | Underestimation | 7 | 23% |
|  | Did not uncover the abdomen | 6 | 19% |
|  | Forgot the value after finishing to count | 5 | 16% |
|  | Did not measure over a full minute | 4 | 13% |
|  | Child too agitated | 3 | 10% |
|  | Did not reset the timer before starting to count | 3 | 10% |
|  | Overestimation | 2 | 6% |
|  | Child standing instead of sitting or lying down | 2 | 6% |

\* Measured with the oximeter rather than manually
MUAC = mid-upper arm circumference measured with a ribbon; SpO<sub>2</sub> = Blood oxygen saturation
measured with an oximeter

#### Temperature

In the retrospective analysis, 3 HCs exhibited either mono- or multi-repeater type of patterns (Table 3), with the most frequent values being 36°C, 36.5°C, and 37°C (Fig S4). In the observational study, the temperature measurement had the lowest skip rate of all signs (Fig 1). In the few instances where the temperature measurement was skipped, HWs primarily cited forgetfulness as the reason. When the temperature was measured, most of the time it was deemed of insufficient quality (Fig 1), with the predominant reasons being inadequate location (e.g. on the chest instead of forehead) or distance from the skin (for infrared thermometers) (Table 4).

#### MUAC

In the retrospective analysis, 7 HCs exhibited either skipper, wrongly evaluated, mono- or multi-repeater type of patterns (Table 3). Often more than one type of pattern was present per HC. The most frequent values were 14, 15, 16 and 17 cm (Fig S4). The observational study found that MUAC measurement, like temperature, was rarely skipped (Fig 1), with the main reasons for skipping being considering the measurement unnecessary for the child’s condition, considering the child too old, or asking the caregiver for the child’s health card to retrieve a MUAC measurement performed in the latest previous consultation (Table 4). MUAC had the highest rate of incorrect values (Fig 1), with the vast majority being done at a wrong place (other than at the mid-upper arm distance) (Table 4).

#### Weight and height

In the retrospective analysis, no skipper behavior patterns were identified for weight, whereas for height 6 HCs exhibited skipper type of patterns. However, 6 HCs exhibited wrongly evaluated type of pattern, primarily based on weight-for-age and weight-for-height z-scores analyses (Table 3). In the observational study, weight and height were both rarely skipped (Fig 1). However, when omitted, it was because HWs considered them irrelevant to the child’s complaint, or relied on parent-reported weight, or visually estimated height without actual measurement. When weight or height was measured incorrectly, it was mainly because of failure to undress the child; often weight was taken while the child wore heavy clothes and/or shoes and height while the child wore shoes or a hat (Table 4). For weight measurements, additional issues included partial weight application on the balance and problems with balance calibration. For height measurements, issues included incorrect positioning on the measurement device, such as heels, shoulders, or head not touching the flat surface, and child not looking straight ahead during measurement (Table 4).

#### Heart rate and blood oxygen saturation measured using an oximeter

In the retrospective analysis, inadequate patterns of heart rate measurement were identified in 8 HCs, predominantly as skipper (5 HCs), but also as mono-(3 HCs) and multi-(2 HC) repeaters (Table 3). The most frequent values were 68, 96 and 99 beats/min (Fig S4). For blood oxygen saturation, inadequate type of patterns were identified in 7 HCs, manifesting as skipper (4 HCs), mono-(3 HCs), multi-(3 HCs) repeaters, and wrongly evaluated (1 HC). The most frequently frequent values were 96, 98, and 99 (Fig S4). The observational study found that heart rate and blood oxygen saturation were indeed frequently skipped (Fig 1) due to time constraints, perceived absence of signs of respiratory distress, and dysfunctional equipment (Table 4). Additionally, incorrectly performed measurements were primarily attributed to the use of inadequate pulse oximeter probes and improper probe positioning (Table 4).

#### Respiratory rate

In the retrospective analysis, respiratory rate was the most problematic sign, with patterns identified in 9 HCs, predominantly as mono-repeaters (4 HCs), but also as skippers and wrongly evaluated (3 HCs each) and multi-repeaters (2 HCs) (Table 3). In the observational study, respiratory rate measurement was the most frequently skipped sign (Fig 1). Justifications included the perceived absence of respiratory distress, time constraints, the possibility to tick “not feasible” in the app, forgetfulness, or reported lack of timing equipment (despite the fact that the RRate app was installed on all tablets) (Table 4). When respiratory rate was still measured, quality was adequate (Fig 1). Notably, the RRate app was used as frequently as the timer (39 times each), but the former resulted in better quality assessments, with 29 out of 39 measurements (74%) being sufficient compared to 18 out of 39 (46%) for the timer. Mistakes included underestimation or overestimation, mismanagement of measurement timing (e.g. starting the timer a lot before starting to calculate RR, not resetting the timer before starting to count, or not making the measure over a full minute), and failure to uncover the abdomen (Table 4).

## Discussion

In this study, we took advantage of data collected during consultations through the use of a digital CDSA to assess the possibility of the presence of clinical skill gaps and to analyze those gaps among HWs working in primary care centers. This exploratory study revealed notable deficiencies in the quality of measurements and assessments among HWs using the ePOCT+ tool in Rwanda. All signs we analyzed had gaps, some predominantly because they were skipped (respiratory rate, blood oxygen saturation, heart rate) and others predominantly because the quality of measurement was insufficient (temperature, MUAC, weight, height).

### Key challenges with vital signs and basic measurements

#### Perceived relevance of measurement

Our results underscore a significant issue with the perceived relevance of certain measurements. Despite the ePOCT+ prompting signs such as heart rate and blood oxygen saturation based on specific criteria related to the child’s condition, these measurements were frequently skipped. Notably, these signs, along with respiratory rate, were among the most frequently omitted. This may be attributed to the lack of integration of pulse oximetry into the Rwanda IMCI guidelines (30), making them only familiar to the HWs in the context of the DYNAMIC study and potentially leading to their misperception as unnecessary. Additionally, despite the availability of a ‘not feasible’ option, the retrospective analysis suggested that some HWs entered a value without actually taking the measurement, possibly to avoid having to explain their decision not to measure during the monitoring visits. This behavior was also observed, though less often, during our observational study, potentially due to the Hawthorne bias. Accurate measurement of respiratory rate is particularly crucial as it can help identify critical signs of respiratory distress and differentiate between mild and severe respiratory infections, which can significantly impact clinical outcomes.

To address this, targeted training should emphasize the importance of each measurement and provide practical strategies for integrating them effectively into consultations. Additionally, to improve the perceived relevance of these measurements, the contextual alerts within ePOCT+ can be revised to briefly explain why specific measurements are necessary based on the child’s symptoms and provide supporting evidence. Furthermore, if a measurement is skipped, HWs could be required to enter a justification, ensuring they reconsider the importance of the measurement and its relevance to the child’s condition. However, this approach should be considered with caution, as making the opting out option more difficult for HW could further encourage entry of false values without taking the measurement.

#### Training gaps and supervision

Regular assessment of basic measurements such as temperature, weight, height and MUAC is part of a primary clinical assessment, regardless of the child’s complaint. In the post-pandemic period in Rwanda, there has been an emphasis on creating triage-like stations in the patient registration area where these measurements are commonly assessed –these stations were present in most (15 of 16) studied HCs but most frequently staffed by non-clinicians. As a result, the vital signs and basic measurement were assessed most of the time in these stations; however the quality of their measurement was compromised due to methodological errors. While task-shifting remains a valuable strategy, it is crucial to ensure that staff who take on these tasks receive high-quality training and ongoing supervision. Training should focus on a specific set of signs measured at the triage station, emphasizing the quality of these essential measurements over the quantity of tasks performed.

To further enhance measurement accuracy, the tablet-based application used in the triage stations includes warnings for numerical values that fall outside acceptable ranges. However, triage station staff who took the measurements wrote them on paper and nurses subsequently entered the values into the tablet in the consultation rooms. As such, any possible alerts incorporated in to ePOCT+ were not available to the staff taking the measurements. Secondly, variations in inadequate measurements by HWs were sometimes insufficient to trigger some alerts. This was highlighted by the retrospective analysis, which revealed extreme values in Z-scores (MUAC-for-age, weight-for-age, and weight-for-height) that did not activate the alert thresholds. Therefore, the algorithm’s alerts may need adjustment to account for Z-scores, not just raw MUAC, height, or weight values.

#### Equipment related challenges

New or less familiar equipment has also presented significant challenges. The introduction of infrared thermometers in HCs during the COVID-19 pandemic was an important initiative aimed at saving time for HWs. However, due to the crisis situation, there may have been insufficient training and guidelines provided for the correct use of these new thermometers, especially to non-clinicians staffing the triage stations. Similarly, the addition of pulse oximeters as part of the DYNAMIC study introduced SpO_2_ and heart rate measurements, which were not traditionally part of primary care assessments. Despite the training provided on pulse oximeter use, issues with its application persisted, potentially due to the high turnover rate of HWs in HCs. Although discouraged by the study team, new staff may have worked with equipment for which they have not received sufficient training.

To address these challenges, a dedicated person at the overseeing district hospital responsible for monitoring and supervision of nurses in HCs could be provided with access to a dashboard with similar comparisons that were explored in the retrospective analysis. When issues with measurements are identified using the data patterns, nurses in that particular HC could be pointed to an instructional video or in-person hands-on refresher training on the use of the tools.

#### Time constraints

Time constraints also emerged as a significant factor affecting the quality of measurements. Respiratory rate, in particular, was the most frequently skipped measurement, with time pressure often cited as a primary justification, even though the RRate app was introduced and available to facilitate its accurate and efficient measurement (i.e., to measure respiratory rate with a timer requires a full minute, whereas the RRate app can determine it accurately in approximately 15 seconds).

While task-shifting has proven effective in redistributing responsibilities, the key challenge remains to ensure that time constraints do not compromise the quality of clinical assessments. One approach could involve increasing the number of triage measurements collected at the reception area based on the patient’s complaint, allowing HWs more time during consultations to focus on more complex assessments. Of course, this would require ensuring that the triage staff receive proper training to perform these measurements accurately and consistently, enabling them to make adequate and reliable assessments at the reception stage. Additionally, streamlining procedures to better integrate time-saving tools, such as ensuring the use of the RRate app, can further alleviate time pressures. Enhancing training on these efficient measurement techniques is also crucial, ensuring HWs are both confident and proficient in using tools that save time without sacrificing accuracy. To reinforce the importance of complete and accurate measurements under time constraints, providing recognition or incentives for HWs who consistently perform high-quality assessments can help sustain motivation and improve overall performance.

### Strengths and limitations

One of the key strengths of our study was the availability of a large and comprehensive consultation dataset and the ability to explore data patterns of clinical signs in detail. Analyzing such extensive data deepened our understanding of healthcare practices, in particular while using digital CDSAs. Our study design (selecting HCs with suspected low clinical performance from the data trends to then observe consultations in these specific HCs to identify the main sources of problems) could be applied not only in various research settings but also by district health management teams in routine conditions. It allows monitoring HWs’ progress over time and providing insights into their performance and skill development through mentorship sessions based on their own consultations data. One example of the particular utility of the data analysis exercise is for weight, height and MUAC measurements. In the retrospective data analysis, measurement quality issues were flagged in the distribution of z-scores (Table 3), but from the raw measurements it was not possible to determine what the specific problems were. Quality issues were subsequently confirmed in the prospective observations, highlighting the use of retrospective analysis as a first indication of a problem.

The study also had several limitations. First, we used the average of all HCs as the reference, due to the lack of a gold standard (which is very difficult to set up in the absence of a large number of experienced healthcare providers to re-examine each child and may anyway exhibit too much variability to be reliable). This approach is justifiable from a practical standpoint of first and foremost identifying the “worst-performing” HCs, as those that deviate most from the average. Second, stemming from the first limitation, we only observed those potentially “worst-performing” HCs, meaning that our findings are not representative of the overall healthcare quality in the study area. Third, the observational study is affected by the Hawthorne bias, meaning that HWs may have changed their behavior due to their awareness of being observed (31). We tried to minimize this bias by emphasizing non-interference. Despite this potential bias, many gaps were observed, suggesting that the Hawthorne effect was likely minimal. Fourth, while individual logins to the CDSA application were provided to all HW and individual-level data were theoretically available, we could not be sure that all HWs used their own login, and thus chose to conduct the analysis at HC level instead. Further analyses and visualizations could be done at individual HW level. Fifth, we did not use the most widely recognized methodology for observing consultations with children, which is the Service Provision Assessment (SPA)(32). This choice was made because the SPA is designed to evaluate adherence to the IMCI guidelines. However, the ePOCT+ protocol included a broader range of pathologies and wider age range of patients, and our objective extended beyond conventional adherence (i.e., whether or not the assessment was performed), aiming to also assess the quality of clinical evaluations conducted. Lastly, we did not formally correlate the findings of the retrospective data analysis and the prospective observations due to the temporal gap between the two parts of the study and potentially different HWs involved. Despite these limitations, the findings of the two parts of the study are generally congruent.

## Conclusions

Although our findings from the observation study may be biased due to selection, the results from the retrospective study are indicative of the overall population of HWs in the study area, revealing deficiencies in clinical skills among the users of the CDSA. Importantly, while we assessed relatively simple parameters and found significant problems, more complex clinical assessments (e.g., palmar and conjunctival pallor, skin pinch, eye examination, ear examination, etc.) could suffer from even more significant gaps (the latter being binary variables, making it difficult to ground truth through retrospective data analysis). Nonetheless, we can conclude that additional training is necessary in order to improve the clinical skills and thus use CDSA to its full potential. Rigorous assessment of HWs’ knowledge and skills is essential before deploying clinical decision support systems. Likewise, encouraging HWs to avoid entering data without performing the necessary exams or measurements would ensure accurate data input and avoid erroneous outputs (21). Strategies for skill improvement, such as on-site personalized mentoring based on HWs’ own data and direct consultation observations, as well as eLearning programs, are crucial. The best approach for skill improvement is clearly on-site training and mentorship, as seen in developed countries through the ‘companionship’ model, where strong and regular supervision of consultations helps maintain high-quality skills (33,34). Unfortunately, this model is largely absent at the primary care level in Africa, representing a significant challenge for improving clinical performance. In this context, eLearning emerges as a promising alternative in Africa, but it comes with limitations. eLearning programs have often yielded results below expectations, as their success depends on fulfilling key conditions, such as active engagement, adequate technological support, and integration with hands-on practice (35–37). Our analysis highlights the potential for continuous monitoring of data trends, allowing to track changes in clinical skills over time and assess the impact of interventions, such as the introduction of eLearning programs. This dynamic approach provides valuable insights into the evolution of HW skills and the effectiveness of training initiatives.

## Data Availability

All database files are available in the Unisanté public data repository (DOI: https://doi.org/10.16909/dataset/55). The database will be made open access upon the acceptance of the corresponding paper. Before the paper's acceptance, access to the database can be requested by emailing.

## Acknowledgments

We extend our sincere gratitude to the DYNAMIC project teams in Switzerland and Rwanda for their indispensable support. Special thanks to the DYNAMIC Rwanda field team for their dedication, and to all participating healthcare workers whose contributions were crucial to this study’s success. We also gratefully acknowledge the Swiss Agency for Development and Cooperation for their financial support (project # 7F-10361.01.01).

